# Distribution of Time Documented as Spent on Observation Unit Disposition Process

**DOI:** 10.1101/2024.12.10.24317182

**Authors:** Farrah Edmond, Nicole Algarin Mojica, Samantha Horowitz, Htet Ng, Matthew Bacchus, Tarek Harharsh, Mark Richman

**Affiliations:** Northwell Long Island Jewish Medical Center, Dept. of Emergency Medicine, 270-05 76^th^ Ave., New Hyde Park, NY 11040

## Abstract

**Introduction:** Emergency Department (ED) Observation Units (OUs) are increasingly-common. Centers for Medicare and Medicaid Services (CMS) reimbursement policies incentivize documenting longer times spent on the disposition/discharge process (≥31 minutes). In addition, there is a well-recognized phenomenon of “terminal digit preference (ending times in 0 or 5). The Long Island Jewish Medical Center (LIJ) ED’s OU disposition process is performed by physician assistants (PAs). This study investigates the distribution of times documented by OU PAs as being spent on disposition and whether those times indicate terminal digit preference.

**Methods:** Documentation for discharge times of 102 patients dispositioned from the LIJ ED OU were captured, along with the PA involved. Students T-test was used to compare percentages. Statistical significance was set a priori at p <0.05.

**Results:** Seven PAs entered discharge times for 102 patients. No times were documented as <31 minutes. Time documented by PAs demonstrated significant clustering (ie, each PA was very consistent in the amount of time they documented). Nearly 50% of times were documented as “35 minutes.” Almost the entire remainder of times (46.1%) were documented as exactly 31 minutes or within 2 minutes of that (ie, 31, 32, or 33 minutes). Slightly over 50% of times documented ended in a “0” or a “5,” statistically-significantly greater than the expected 20% (p <0.0001).

**Discussion:** Documentation of most times being at or just slightly greater than 31 minutes suggests that documentation may reflect CMS’s reimbursement incentives. Over half of times documented ended in 0 or 5, indicating “terminal digit preference.” Further research is needed to confirm these findings and analyze the impact of CMS policy changes on in healthcare documentation.

## Introduction

Following initial evaluation, Emergency Department (ED) patients are discharged, admitted, or placed into an observation unit (OU) for further assessment and treatment. OUs use has been increasing over the past 25 years,^i^ as they are associated with decreased length of stay, costs, and increased patient satisfaction.^ii^

The Centers for Medicare and Medicaid Services (CMS) reimburses providers for dispositions or care based on time in several scenarios. Qualification for OU billing and reimbursement is proportional to time spent on a case, including the disposition (admission or discharge) process; this is consistent with how Medicare reimburses for other situations, such as critical care.^**iii**^ Setting reimbursement rates proportional to time spent on a case has been associated with “upcoding” of time (ie, documenting more time on a case than was spent).^iv^

In addition, terminal digit preference is a well-known phenomenon in which persons preferentially round a number up or down toward a preferred, commonly-used digit (eg, 0 or 5). This has been described in reporting age,^v^ blood pressure,^vi^ and tumor size.^vii^ Such a phenomenon might also apply to documenting time spent on tasks.. If times were documented according to true statistical distribution, one would expect the terminal digit of the time documented to be evenly-distributed (eg, 10% of times documented would end in “1,” 10% of times documented would end in “2,” etc.)

The Long Island Jewish Medical Center ED staffs a 12-bed OU with Attending Physician and Physician Assistant (PA) coverage. The PA is responsible for documenting notes, with the Attending largely signing off on the PA’s documentation. In late 2023, ED administration notified faculty and PAs that CMS reimburses at a higher rate when time spent on disposition is documented at >31 minutes. We hypothesized this very specific time would encourage PAs to document time spent on disposition closely-above 31 minutes. Furthermore, terminal digit preference raises the possibility PAs may preferentially document time spent as ending in “0” or “5.” We hypothesize the percent of times documented that end in “0” or “5” will be disproportionate to their expected percent were they truly randomly distributed; we expect to see, for example, 25% of times end in “0” and 30% of times end in “5,” whereas, were the times truly randomly distributed, 10% of times would end in “1,” 10% should end in “2,” etc.

This study aims to determine the distribution of time documented as being spent on the disposition process from LIJ ED’s observation unit.

## Methods

Northwell Health is a 22-hospital health system largely operating in Long Island and New York City. Long Island Jewish Medical Center is a 583-bed tertiary care teaching hospital serving an ethnically and socio-economically diverse population. The adult Emergency Department (ED) sees approximately 100,000 patients per year. The ED staffs (by an Attending physician and a PA) an OU with an annual census of ∼3,000 patients. The OU has 12 beds; occasionally, when more than 12 patients qualify for observation status, up to two additional patients are put in “observation status” while physically in another part of the ED (“virtual observation”). Patients managed in the OU typically have these conditions: chest pain (for cardiac stress test and/or echocardiogram); mild-moderate asthma; pharyngitis or peritonsillar abscess; transient ischemic attack, minor stroke, or significant back pain who need MRI (which takes a long time in the queue and the machine, and for interpretation); and significant anemia requiring blood transfusion (which takes 3 hours per unit transfused). OU patients are usually kept for 24-36 hours. When OU patients’ admission or discharge disposition has been decided on the basis of clinical status, laboratory and radiology results, and consultation (eg, Ear, Nose, and Throat), the observation unit PAs allocate time to the disposition process and document time spent on this process.

For this study, we retrospectively collected data from 102 patients who were in the OU between June 1, 2024 and July 31, 2024. Patients were included if they were dispositioned from LIJ ED’s OU. No patients were excluded from the study, as there was no exclusion criteria. Patient data was included on a convenience sample basis (when the Principal Investigator was in the ED to supervise research assistants). Data collected included the amount of time documented as having been spent on the disposition process, the name of the PA (to evaluate for clustering (eg, whether the same PA documents the same amount of time for each patient)), and the disposition diagnosis. The 7 PAs were de-identified by replacing names with numerical values ranging from 1 to 7. Sorting the patients by PA and including the disposition diagnosis allowed for analysis of time documentation patterns by individual PAs.

Univariate statistical analysis was performed on time data. Specifically, we calculated the average time and standard deviation documented as having been spent in the disposition process; the percent of times within 5 minutes more or less than 31 minutes; and the percent of times that end in “0” or “5.” Furthermore, we investigated distribution of time documented by each particular PA: for each PA, the average time and standard deviation was documented; the percent of times a particular time was documented (eg, for PA #1, the percent of times documented as “31,” “32,” 35,” etc.); the percent of times less than 31 minutes; and the percent of times that end in”0 “ or”5.” Statistical analyses and graphs were performed using Microsoft Excel 2021 (Microsoft Corporation, Redmond, WA). Students T-test was used to compare percentages. Statistical significance was set a priori at p <0.05.

This study was deemed exempt by Northwell Health’s Institutional Review Board (IRB #: 24-0624).

## Results

Seven PAs entered discharge times for 102 patients. No times were documented as under 31 minutes. Average times documented by PA varied between 31 and 37 minutes. (**Figure 1**) Time documented by PAs demonstrated significant clustering (ie, each PA was very consistent in the amount of time they documented). (**Table 1**) Nearly 50% of times were documented as “35 minutes.” Almost the entire remainder of times (46.1%) were documented as exactly 31 minutes or within 2 minutes of that (ie, 31, 32, or 33 minutes). (**Figure 2**)

**Figure 1.**
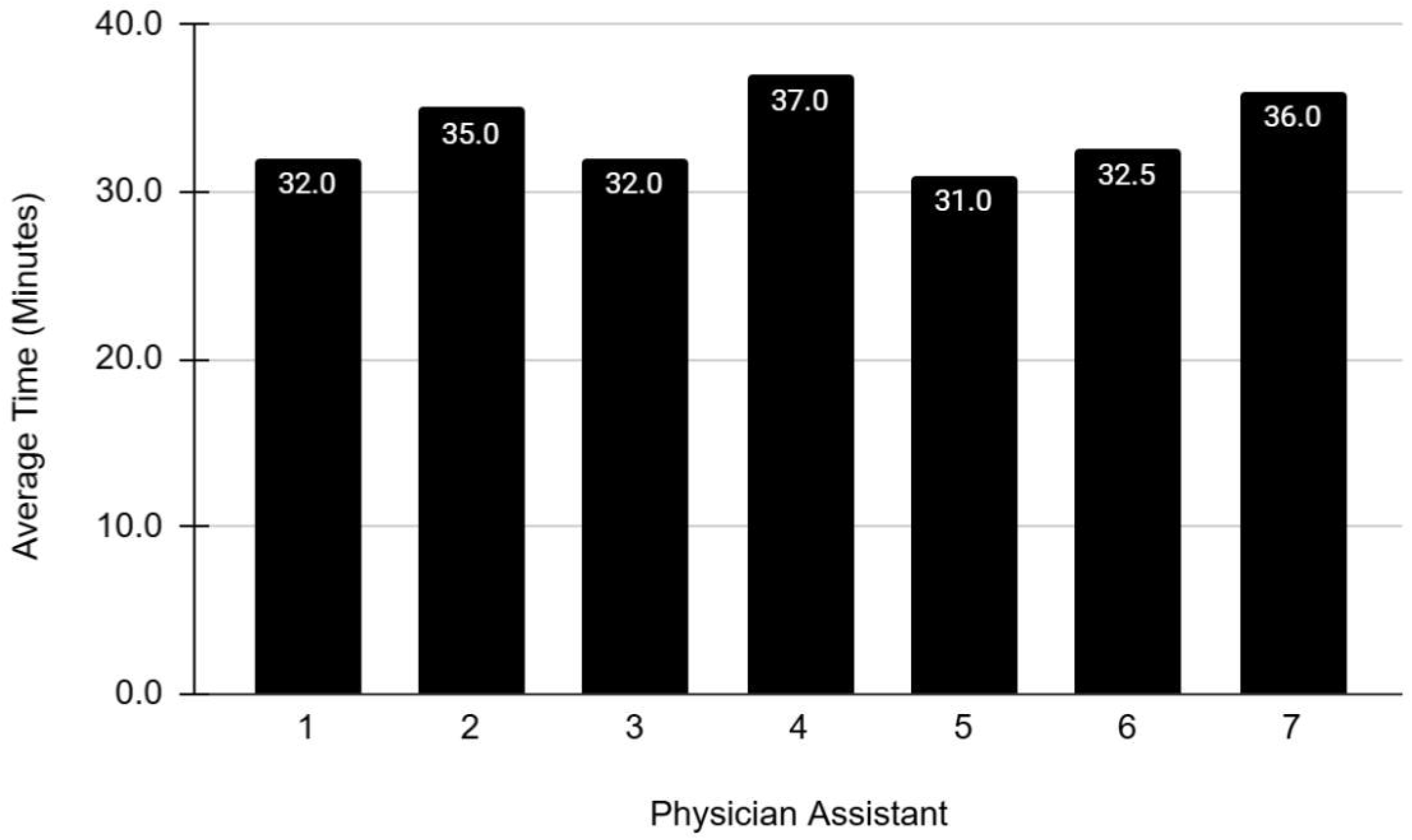

**Table 1.**

| PA | Minutes documented (# of patients with this documentation time) |  |  | Total # of patients | # of different times documented | Comments |
| --- | --- | --- | --- | --- | --- | --- |
| PA 1 | 32 (14) | 33 (14) |  | 28 | 2 | 50% documented as 32 minutes, 50% as 33 minutes |
| PA 2 | 32 (3) |  |  | 3 | 1 | 100% documented as 32 minutes |
| PA 3 | 34 (1) | 35 (41) | 40 (2) | 44 | 3 | 93% documented as 35 minutes |
| PA 4 | 32 (10) |  |  | 10 | 1 | 100% documented as 32 minutes |
| PA 5 | 35 (8) | 45 (2) |  | 10 | 2 | 80% documented as 35 minutes |
| PA 6 | 31 (6) |  |  | 6 | 1 | 100% documented as 31 minutes |
| PA 7 | 36 (1) |  |  | 1 | 1 | 100% documented as 36 minutes |

**Figure 2.**
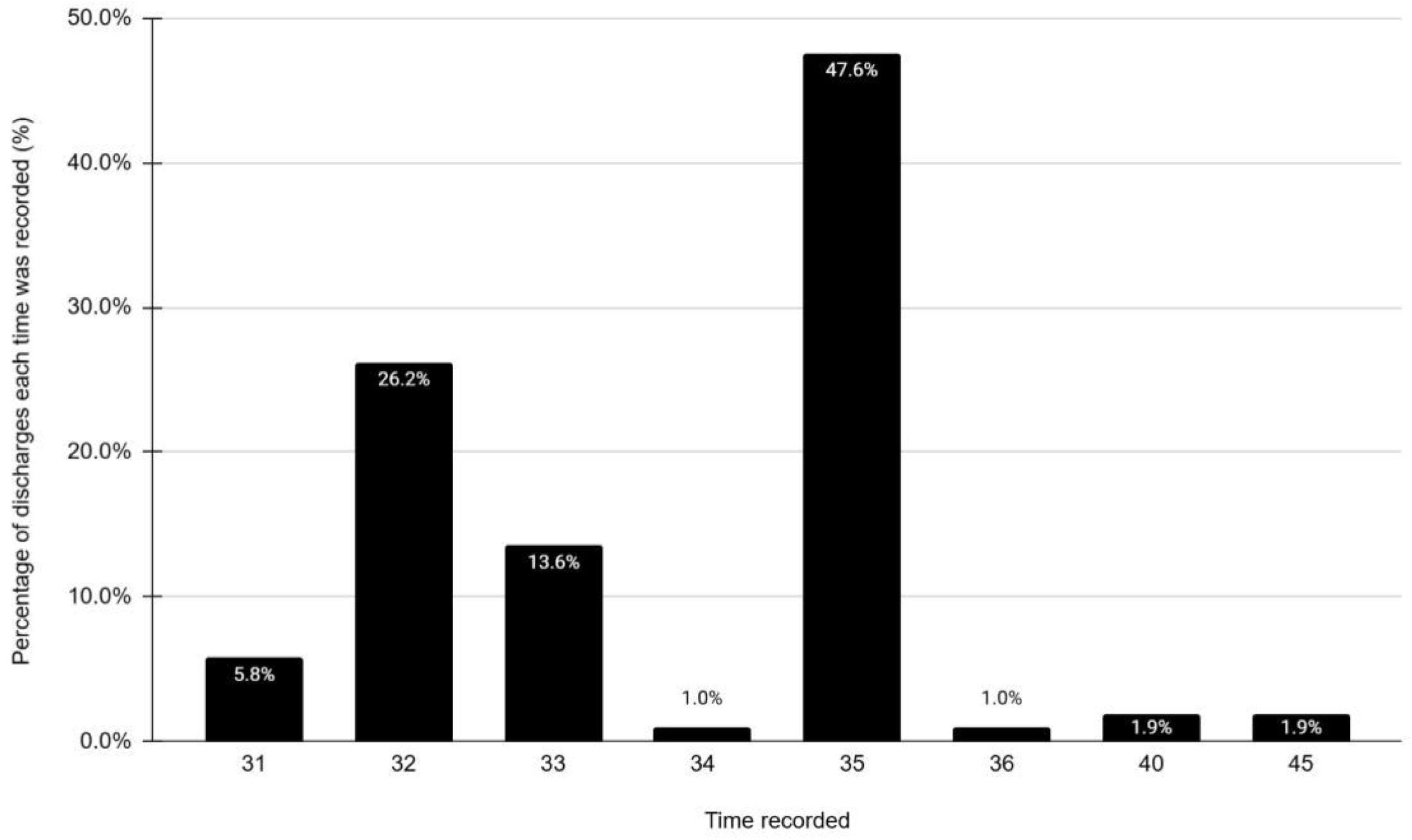

Slightly over 50% of times documented ended in a “0” or a “5,” well beyond the 20% of total time one would expect if documentation reflected the expected random distribution (eg, 10% of times should end in “1” such as 41 minutes, 10% should end in “2” such as 42 minutes, etc.). (**Figure 3**) The 51.4% of dispositions whose times were documented as ending in “0” or “5” was statistically-significantly greater than the expected 20% (p <0.0001).

**Figure 3.**
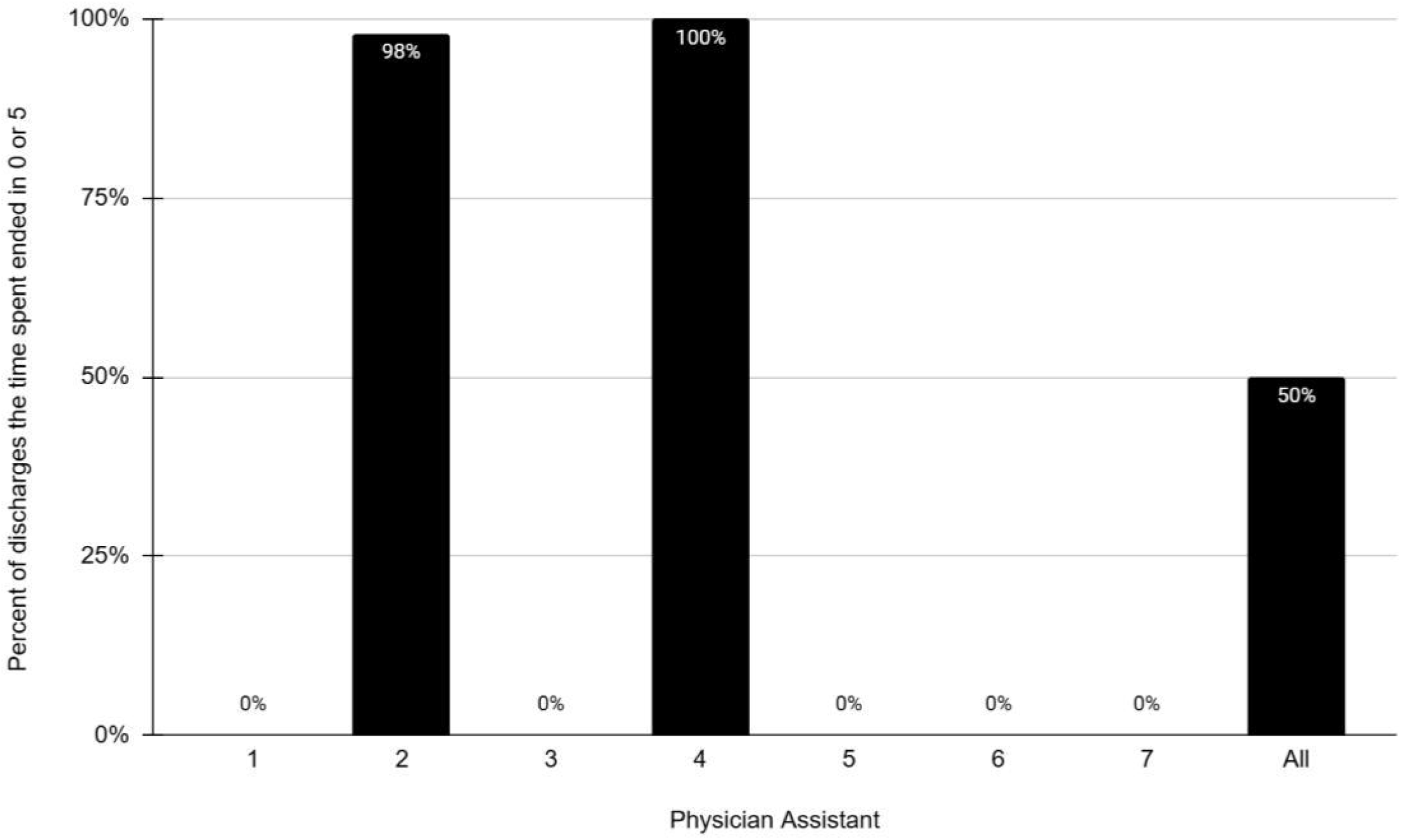

## Discussion

PAs documented a time spent on discharge/disposition activities of 31 minutes or more for all 102 patients seen in the OU, recording a number ending in 0 or 5 approximately 50% of the time. Such consistency suggests PAs may be trying to meet CMS’s OU reimbursement incentives, and following terminal digit preference in their time documentation.

Previous literature suggests incentives may encourage providers and healthcare organizations to “game the system” to, on paper, meet the incentives, without actually performing the necessary steps or achieving the goals.^viii^ Authorities in behavioral economics and medical quality of care^ix^ suggest incentives must closely align to professional values.^x^

Inaccurate documentation, reflected as errors in the medical record, is commonplace. One study of 105 encounters covertly recorded by audio of 36 physicians found 636 documentation errors (181 errors of commission (entering findings that did not take place) and 455 errors of omission (not entering important findings that were found). Ninety percent of notes contained at least one error. In 21 (20%) of notes, the charting resulted in a higher billing level than the audio recording.^xi^

This study had several limitations. First, this was a single-site study; OU time documentation practices may differ at other institutions. Second, as an exploratory study, there was a small sample size of both patients and PAs. Nonetheless, this research preliminarily confirms our hypothesis that, given CMS’s reimbursement policy (higher rates for >31 minutes), the majority of times documented would be slightly above 31, especially 35 or 40, owing to terminal digit preference. Third, we performed only a univariate analysis, not adjusting for potential confounding variables such as diagnosis, need for translation services, or complexity of the ED or OU visit (eg, need to communicate with consultants regarding discharge plans). We will follow these observations with a larger study that includes multivariate analysis. Finally, we did not follow the PAs around during their discharge process to determine the amount of time they spent on the process or its documentation, and, therefore, whether the amount of time they documented was, indeed, accurate.

## Conclusion

The consistency among reported times (46% documented at 31, 32, or 33 minutes), and the disproportionate representation of times ending in “0” or “5”, in this exploratory research project suggests OU providers inaccurately report time spent in the process and documentation of discharging OU patients. The next steps in our research will be to determine what percent of times documented being spent on discharge were ≤31 minutes prior to the late 2023 announcement of better reimbursement for times >31 minutes. Evidence of a substantial change in documented times from being frequently below 31 minutes to none being below 31 minutes would add weight to the observation that OU providers have adapted their documentation practices in response to the time-based incentive.

## Data Availability

The data is available upon request from the corresponding author, Dr. Richman. The data utilized for this research were provided by Dr. Richman, who works in the Emergency Department at LIJ, through his portal access.

## References

i Ross MA, Granovsky M. History, Principles, and Policies of Observation Medicine. Emerg Med Clin North Am. 2017 Aug;35(3):503–518. doi: 10.1016/j.emc.2017.03.001. PMID: 28711121.

ii American College of Emergency Physicians. Emergency Department Observation Unit Utilization Trends From 2006 to 2019 (1/23/24). Available at: https://www.acep.org/observation/newsroom/jan-2024/emergency-department-observation-unit-utilization-trends-from-2006-to-2019. Accessed 7/3/24.

iii Medicare. Medicare Evaluation and Management Services Guide. Available at: https://www.cms.gov/outreach-and-education/medicare-learning-network-mln/mlnproducts/downloads/eval-mgmt-serv-guide-icn006764.pdf. Accessed 6/30/24.

iv Coustasse A, Layton W, Nelson L, Walker V. Upcoding Medicare: Is Healthcare Fraud and Abuse Increasing. Perspect Health Inf Manag. 2021 Oct 1;18(4):1f. PMID: 34975355; PMCID: PMC8649706.

v Denic S, Khatib F, Saadi H. Quality of age data in patients from developing countries. J Public Health (Oxf). 2004 Jun;26(2):168–71. doi: 10.1093/pubmed/fdh131. PMID: 15284321.

vi Nietert PJ, Wessell AM, Feifer C, Ornstein SM. Effect of terminal digit preference on blood pressure measurement and treatment in primary care. Am J Hypertens. 2006 Feb;19(2):147–52. doi: 10.1016/j.amjhyper.2005.08.016. PMID: 16448884.

vii Tsuruda KM, Hofvind S, Akslen LA, Hoff SR, Veierød MB. Terminal digit preference: a source of measurement error in breast cancer diameter reporting. Acta Oncol 2020;59:260–267.

viii Glasziou PP, Buchan H, Del Mar C, Doust J, Harris M, Knight R, Scott A, Scott IA, Stockwell A. When financial incentives do more good than harm: a checklist. BMJ. 2012 Aug 13;345:e5047. doi: 10.1136/bmj.e5047. PMID: 22893568.

ix Himmelstein DU, Ariely D, Woolhandler S. Pay-for-performance: toxic to quality? Insights from behavioral economics. Int J Health Serv. 2014;44(2):203–14. doi: 10.2190/HS.44.2.a. PMID: 24919299.

x Roland M. Incentives must be closely aligned to professional values. BMJ. 2012 Sep 10;345:e5982. doi: 10.1136/bmj.e5982. PMID: 22963938.

xi Weiner SJ, Wang S, Kelly B, Sharma G, Schwartz A. How accurate is the medical record? A comparison of the physician’s note with a concealed audio recording in unannounced standardized patient encounters. J Am Med Inform Assoc. 2020 May 1;27(5):770–775. doi: 10.1093/jamia/ocaa027. PMID: 32330258; PMCID: PMC7647276.

